# Comparing Waves of COVID-19 in the US: Scale of response changes over time

**DOI:** 10.1101/2022.03.01.22271713

**Authors:** Robert L. Richards, Grant Foster, Bret D. Elderd, Tad A. Dallas

## Abstract

Local response to the SARS-CoV-2 pandemic differed spatially across the United States but the drivers of this spatial variation remain unclear. We approach this open question by studying the relationship between the growth rate of subsequent waves of the pandemic at the county level during the first year of the pandemic, asking whether state or county demographics better explain variation in this relationship. We found clear spatiotemporal patterns in the relationship between the slopes of subsequent waves in a given county. Generally the standardized difference between the growth rates of waves 1 and 2 and waves 2 and 3 were strongly positively correlated over short distances and shifted to a weak negative correlation at intermediate distances. We also found that peer county health group (a categorization of counties by demographic information relevant to public health) explained variation in response better between wave 1 and 2, while state identity was most important between wave 2 and 3. Taken together, we suggest that there are identifiable spatial patterns in pandemic response across the US but that the nature of these patterns change over the course of the pandemic.

## INTRODUCTION

Beginning early in the pandemic of SARS-CoV-2 both epidemic severity and local response to the pandemic differed spatially across the United States [1, 2]. There is a long history of the study of spatial variation in the basic reproductive number of pathogens [1] but drivers of spatial patterns in pandemic response are less well studied. Much work has been done both to predict the effect of a variety of Non-pharmaceutical interventions (NPIs) on future transmission and to measure the actual effect of NPIs implemented during the pandemic [3, 4, 5, 6, 2]. However, determining which underlying factors contribute to the response to subsequent pandemic waves in a particular location has proven challenging, partially due to the haphazard way in which NPIs were implemented and the difficulty of measuring individual compliance with state mandates [7, 8, 9]. Despite these challenges, identifying the key factors associated with good and bad pandemic responses should be both of general interest and useful for targeting of public health initiatives.

Spatio-temporal variation in response to the pandemic likely results from the interaction of two broad categories of factors: one local, county demographics, and the other regional, state policy. State and local governments began implementing mandated NPIs in the middle of March 2020 and first relaxed any restrictions at the end of April 2020, but the timings of implementation and relaxation and the extent of restrictions varied substantially from state to state [2]. The degree to which individuals and communities within a state enforced and complied with existing policies has also clearly varied [8, 9] and therefore may differ spatially due to local demographics. In fact, direct measurement of the effect of specific NPIs has largely relied on this inherent spatial variation in both the timing of policy implementation and the relationship between policy and actual behavior [2, 4]. If the stochastic processes of disease transmission are responsible for more variation than the local or regional factors, then there may even be no pattern in the responses. Here we consider whether state-level policies and local demographic differences are important to determining the response of a county to COVID-19 and if one category is more useful than the other.

Identifying the factors that contribute to especially good or especially bad pandemic response will allow us to predict areas of increased risk of repeated extreme epidemics and to begin to identify which points can be leveraged to improve pandemic response. We focus on the relative response to the pandemic by comparing the relationship between a given COVID-19 wave and the wave immediately prior to it. Here we use the word “wave” to describe a period of time during which most counties in the US experienced a single peak in COVID-19 cases. Counties which tend to have smaller relative outbreaks in a subsequent peak (good responses) may have collectively improved their NPI compliance or introduced other protective behaviors while those with comparable or larger subsequent epidemics (bad responses) may have not improved or regressed in those behaviors. Although it may be interesting to simply identify which particular counties are especially good or bad at responding to subsequent pandemic waves, we also investigated to what extent patterns of response were related to state or county demographics and over what distances spatial patterns in response similarity were detectable. Because of the changing nature of attitudes and policies regarding the ongoing pandemic the relative importance of state policies and county demographics should similarly change over time. We predict that state level factors should increase in importance as time progresses because the relatively uniform policies early in the pandemic quickly broke down into a state-by-state patchwork of wildly different regulations. Similarly, we expect that county-level demographic factors should also increase in influence over time as many states rolled back restrictions and disease control relied on county level policies and behavior patterns. Finally, we predicted that spatial relationships between county responses should begin positive and decay with distance but that the shape of this decay should vary regionally due to differences in the sizes of both counties and states.

## METHODS

### Data

Case data for SARS-CoV-2 in US counties was compiled by the Center for Systems Science and Engineering at Johns Hopkins University [10]. These data were then rescaled to cases per 100,000 residents based on county population estimates from the US Census Bureau and smoothed using a 14 day moving average. We focused our study on three initial waves of infections. The first wave (January-May 2020) contained the first known emergence of SARS-CoV-2 into counties in the US as well as the first introductions of NPIs to slow spread, the second wave (July-September 2020) represents the first major resurgence of the virus across the US as behavioral changes began to fade over the summer, and the third wave (September 2020 - January 2021) represents the final and largest surge of cases prior to wide availability of vaccines. Counties were grouped for further analysis by state and peer county groups based on categorizations from the Community Health Indicators Project which are themselves constructed according to a number of demographic criteria for the purpose of comparing health outcomes [11]. Data on seroprevalence by county were sourced from the blood donor seroprevalence survey and summarized by wave period [12] for use as an independent predictor variable in analyses.

### Defining waves

We used a modified peak detection algorithm to identify county-level peaks of the epidemic curves in each of the 3 initial waves of COVID-19 infections in the United States prior to widespread vaccination and the spread of variants of concern [13]. Briefly, the original algorithm identifies points which are a 2 standard deviations above or below the mean value of a sliding window. We extend this method by (i) identifying the largest positive “peak” in each established wave period as the top of the epidemic curve for that wave (ii) identifying the last “negative” peak prior to the top of the epidemic curve as the location of the preceding valley and (iii) identifying the start of the new wave as the first positive “peak” identified between the preceding valley and the top of the epidemic curve. This first identified “peak” is simply the first detectable positive anomaly which represents the beginning of the new upward trajectory of the epidemic wave. To estimate the severity of each wave in each county we first took the slope of each epidemic curve (height of the curve/time from the start of the curve, as defined in (iii) above, to the top of the curve) and then standardized the natural log of these slopes to *z*-scores within each wave:

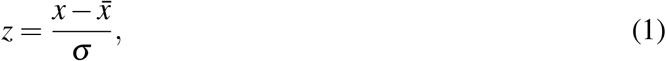

where *x* is the ln(slope) of a wave in a single county, 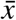 the mean of the ln(slope) across all counties during the relevant wave, and *σ* the standard deviation of those rescaled slopes. If no positive peak was detected during a wave interval then the county was classified as having no wave.

### Statistical analysis

We expected neighboring counties to have more similar wave-to-wave responses than more distant counties. We tested this hypothesis and estimated the distance at which wave-to-wave responses in counties were no longer positively correlated using spatial correlograms constructed with a 100 km interval with the *ncf* package [14, 15]. Spatial correlograms were restricted to the continental United States and separated them by US census region. Prior to visualization, correlograms were trimmed to include only bins with at least 1% of the total number of possible county pairs.

In an effort to understand what factors best explain the variation in wave to wave relationships within counties we fit a suite of mixed effects models to the relationship between subsequent waves and compared their relative fit. We fit a suite of linear models with wave 2 as a dependent variable and wave 1 and estimated seroprevalence during wave 2 as the independent fixed effects. The suite of models included every combination of the random intercept and random slope effects of state and peer county group. Model comparison and selection was then performed using corrected AIC (AICc). All models were fit using the the package *glmmTMB* [16] and we identified the best models using the the *MuMIn* package to compare model AICc [17, 18]. This process was repeated for the relationship between wave 2 and wave 3. Mixed effect model analysis was limited to counties for which seroprevalence data was available.

Data and code to reproduce these analyses are available at https://doi.org/10.6084/m9.figshare.19285379.

## RESULTS

### Spatiotemporal patterns

Peak detection was successful in the majority of counties for all three waves and improved with later waves. Some counties (e.g. rural Utah) proved consistently challenging for successful peak detection (no positive peak detected in a wave) and are therefore excluded from further analysis. The areas of the US with the highest peaks shifted from wave to wave, mostly according to known dynamics of the pandemic. For example, the northeastern US suffered from comparatively severe first waves but comparatively smaller later waves, while in the southeast large waves occurred in spatial pockets in wave 1, the large waves expanded in wave 2, and many areas suffered comparatively smaller waves by wave 3 (Figure 1).

**Figure 1.**
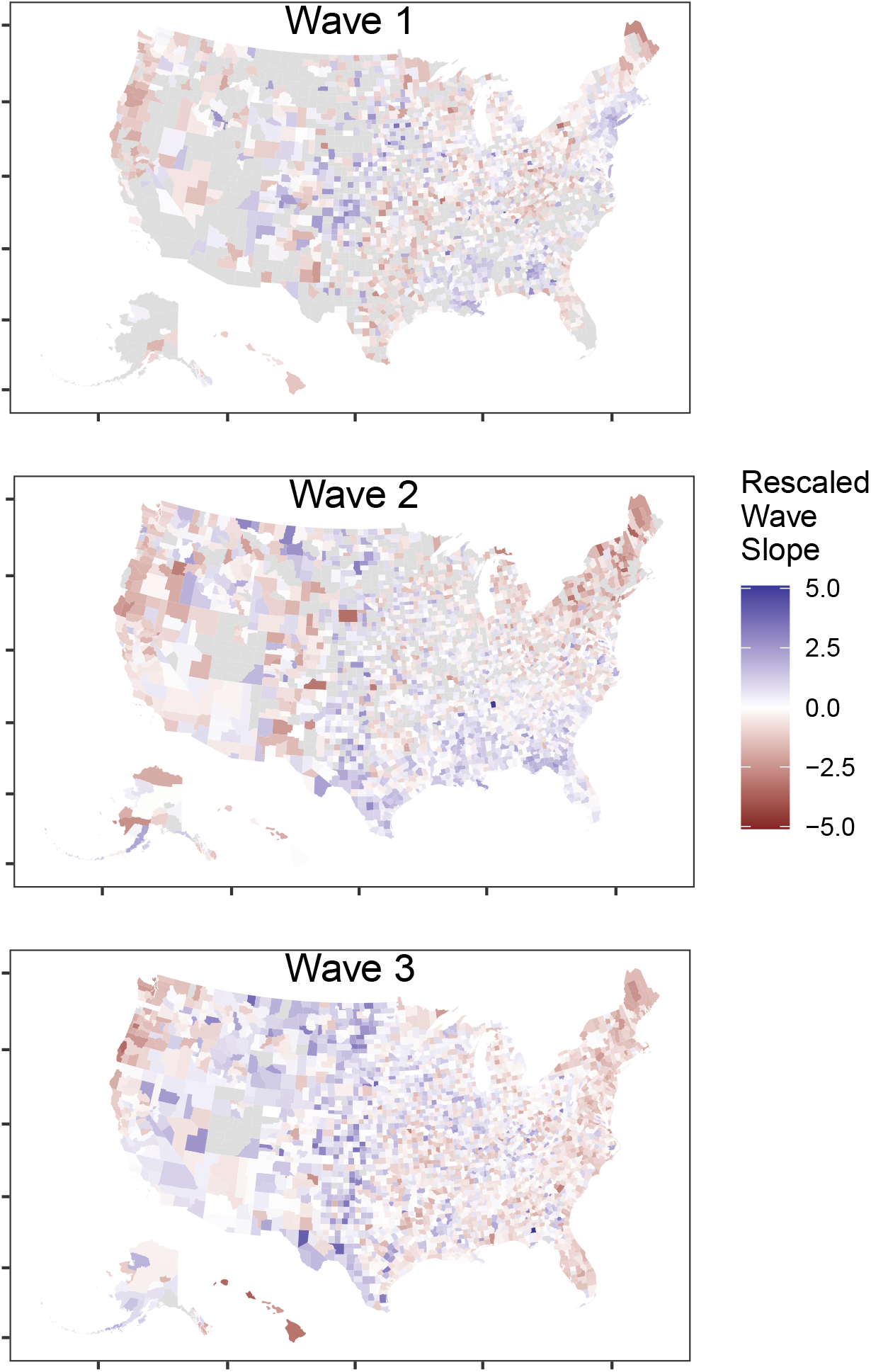
Relative wave slope (z-score) by county for the first 3 waves of COVID-19 in the United States. Many counties lacked large enough epidemics in the first and second waves to detect a clear wave peak. Highest relative wave peaks appear to have shifted from the northeast US (wave 1) to the southeast US (wave 2) to the plains and mountain west (wave 3).

### Wave-to-wave comparisons

We found clear spatiotemporal patterns in the relationship between subsequent waves in a given county (Figure 2). For example, parts of the south and midwest/plains regions of the US have strong positive relationships between waves 1 and 2 (bad resonse; Dark blue-green in Figure 2). While much of the southeast US transitions to a negative relationship between wave 2 and wave 3 (good response; blue), the midwest and plains regions mostly remain high for both later waves (bad response; green and dark blue-green, Figure 2). While both southeastern and plains states tended to have more lax NPI policies the two regions seem to have responded in starkly different ways to repeated COVID-19 waves. These broad regional patterns of similar responses were supported by spatial correlation analyses. The standardized difference between the slopes of both waves 1 and 2 and waves 2 and 3 were strongly positively correlated over short distances, shifting to a weak negative correlation at intermediate distances (Figure 3). Interestingly, the wave 1 to wave 2 transition was characterized both by stronger positive correlations over very short distances and a return to positive correlations over long distances. The scale of these transitions seems to differ slightly by region with correlations in the northeast US decaying the most quickly with distance and the western US displaying significant positive correlations over the longest distances.

**Figure 2.**
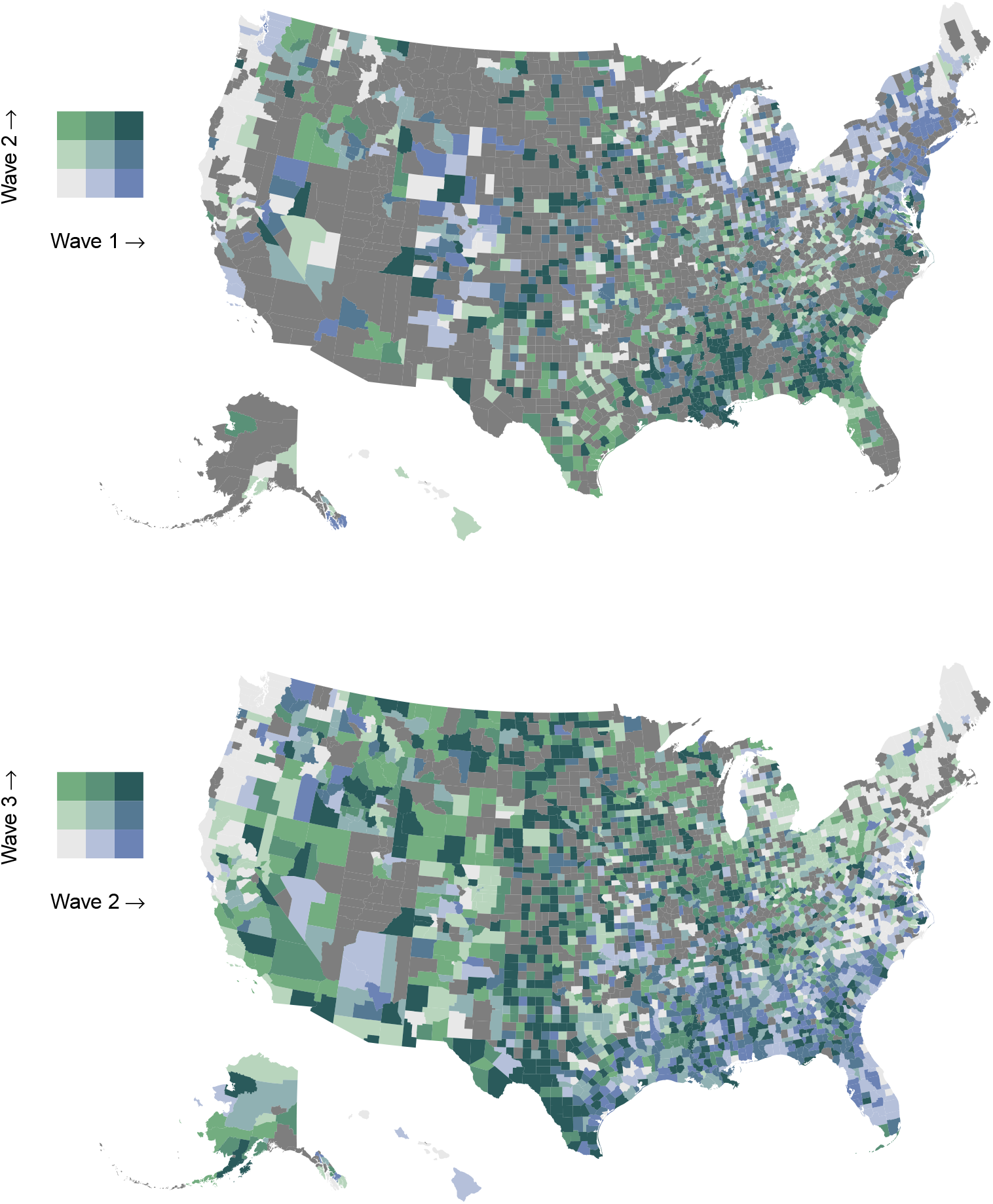
Bivariate map displaying the relationship between the relative slopes of (a) waves 1 and 2 and (b) waves 2 and 3. Deeper blues represent larger prior wave slopes and deeper greens represent larger later wave slopes. Blue areas of the map (the northeast US in wave 1-2 and southeast US in wave 2-3) show improvement in response from wave to wave. While green areas of the map (the southeast US in wave 1-2, and the plains and mountain west in wave 2-3) show regression or stagnation in response from wave to wave.

**Figure 3.**
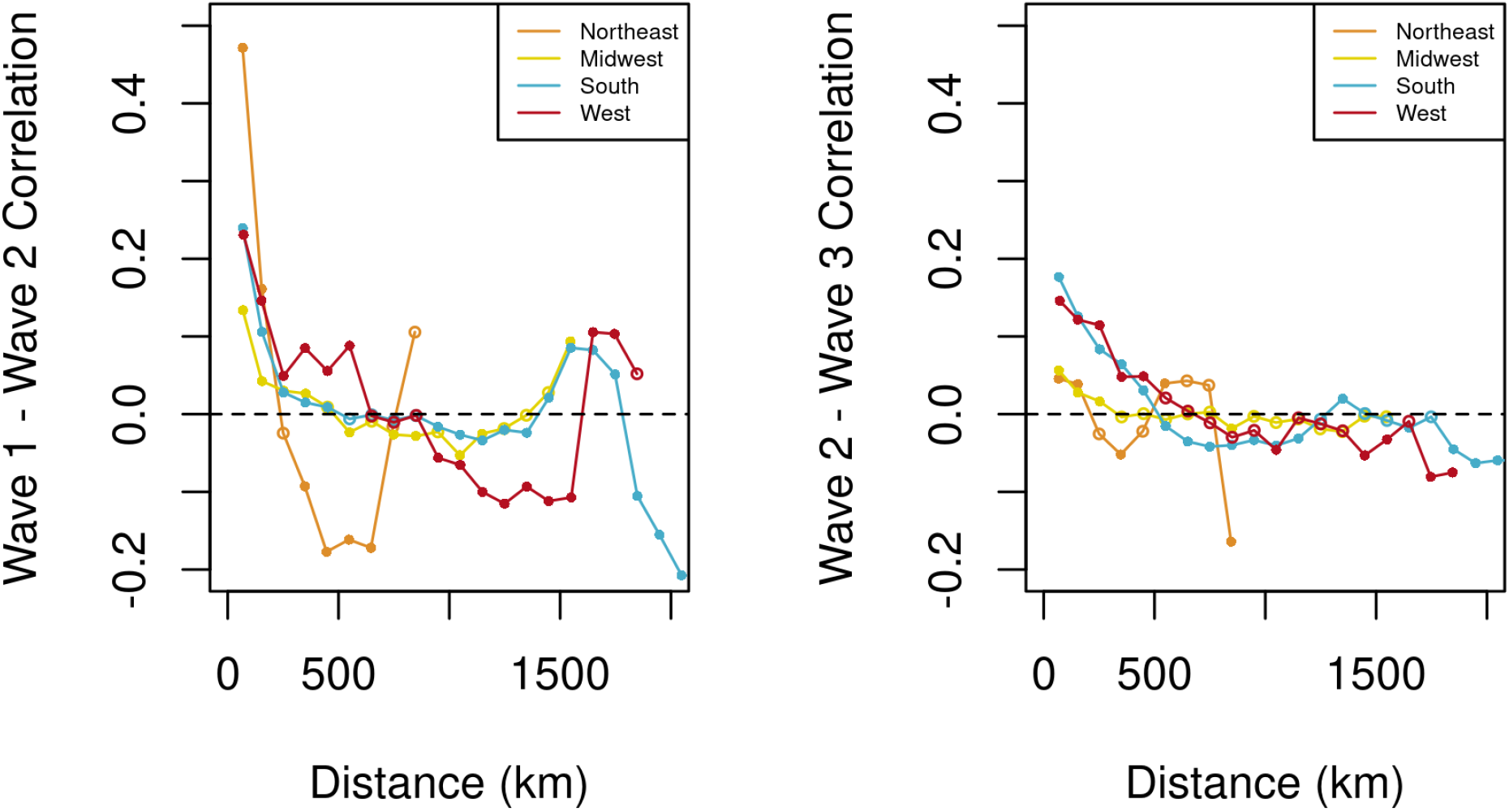
Correlograms show the spatial correlation of the difference between the standardized slopes of subsequent waves by US census region. Both neighboring counties and distant counties are much more strongly correlated in (a) their response from wave 1 to wave 2 than (b) from wave 2 to wave 3, while both figures show neutral or negative correlation at intermediate distances. Closed circles are significantly different from zero while open circles are not.

Using mixed effect models we found that the peer county health group contained slightly more information for wave 1 and wave 2, while the random effect of state contained the most information for fitting the relationship between wave 2 and wave 3. In particular, for waves 1 and 2, the best model was one with a random slope for peer county health group and only the model with no random slopes was comparable (Table 1). Similarly, in the most complex model the variance explained by the random slope of peer county group was significantly different from zero (95% CI: 0.02 - 0.22) but the random slope of state was not (95% CI: 0 - 0.16). For waves 2 and 3, comparison via AICc identified the model including random slopes of state as the best fit and the models with no random slopes and that with random slopes for both state and peer county group were the next ranked models (Table 1). This pattern was reinforced by the fact that in the most complex model the variance explained by the random slope of state was significantly different from zero (95% CI: 0.01 - 0.17) while that explained by the random slope of peer county group was not (95% CI: 0 - 0.17). Contrary to our expectations it seems that state-level factors became more important later in the epidemic while county demographic factors dominated early. For peer county groups (groups of counties which are demographically similar) we find some negative relationships for waves 1 and 2, suggesting that those groups substantially improved their response on average (Figure 4). Notably, the most negative peer county groups (and therefore the best responders) for waves 1 and 2 include groups 1 and 2 which represent high population density urban areas (Table 2). While all states have a positive relationship between subsequent waves, the slope varies substantialy for the wave 2 to wave 3 response.

**Table 1.** Ranking of mixed-effects models predicting a wave slope. All models include the slope of the prior wave and the seroprevalence during the predicted wave. In the table we specify the random effects in each ranked model. Models are ranked by ΔAICc with the number of degrees of freedom (df), and Akaike weights (*w*_*i*_). Only models with ΔAICc ≤ 2 are shown.

| Wave 1 to Wave 2 | df | $\Delta\text{AICc}$ | $w_i$ |
| --- | --- | --- | --- |
| Peer County: Intercept + Slope; State: Intercept | 8 | 0 | 0.619 |
| Peer County: Intercept; State: Intercept | 6 | 1.75 | 0.258 |
| Wave 2 to Wave 3 | df | $\Delta\text{AICc}$ | $w_i$ |
| Peer County: Intercept; State: Intercept + Slope | 8 | 0 | 0.32 |
| Peer County: Intercept + Slope; State: Intercept + Slope | 10 | 0.52 | 0.247 |
| Peer County: Intercept; State: Intercept | 6 | 0.7 | 0.225 |
| Peer County: Intercept + Slope; State: Intercept | 8 | 0.87 | 0.207 |

**Figure 4.**
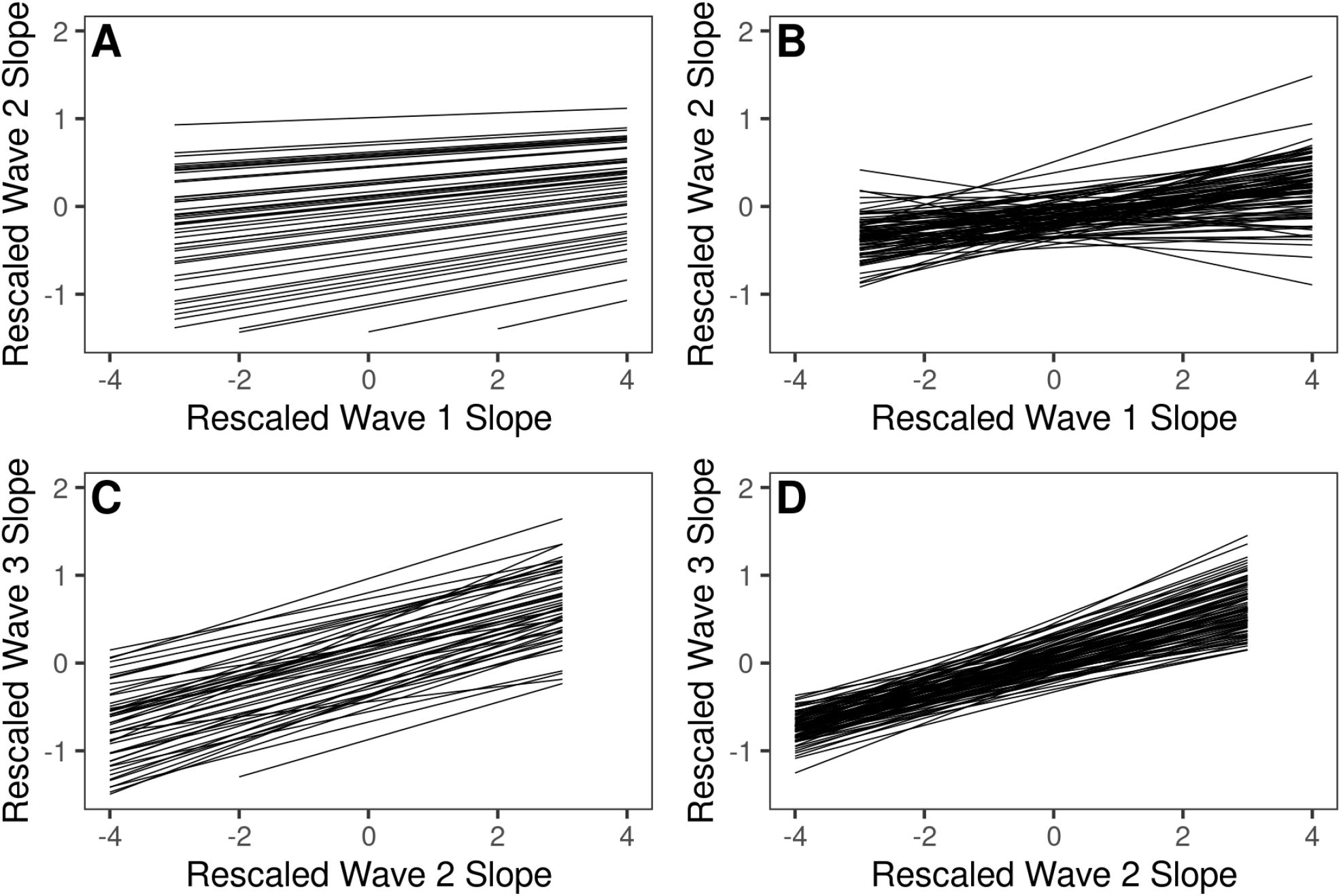
Fitted slopes of the relationship between (a,b) wave 1 and wave 2 and (c,d) wave 2 and wave 3. Slopes are separated by the random effect of (a,c) state and (b,d) peer county group. Fitted relationships are based on the most complex fitted model including all random effects. (b) Peer county group was most important to the relationship between waves 1 and 2, with slopes of the relationship spanning zero, but (c) state was most important to the relationship between waves 2 and 3

**Table 2.** County groups rank ordered by COVID-19 response between subsequent waves according to the best fit mixed effects model. Best learners have the smallest slope between a prior and subsequent wave and the worst learners have the largest slope. For wave 1 to 2 the best fit model identified peer county groups as the most useful random effect. As these peer county groups are not well known we provide a sampling of example counties. Full county lists can be found in the supplementary data.

| Wave 1 to Wave 2 - Best Learners | slope | Wave 2 to Wave 3 - Best Learners | slope |
| --- | --- | --- | --- |
| Group 1 (e.g. LA County, CA; Fulton County, GA; New York County, NY) | -0.185 | Alabama | 0.056 |
| Group 63 (e.g. Alleghany County, VA; Fayette County, IL; Tyler County, TX) | -0.15 | Pennsylvania | 0.084 |
| Group 76 (e.g. Clay County KS; Jackson County, MN; Big Horn County, WY) | -0.103 | Texas | 0.092 |
| Group 2 (e.g. Denver County, CO; Arlington County, VA; Maricopa County, AZ) | -0.066 | Indiana | 0.111 |
| Group 84 (e.g. Clay County, IL; Butte County, SD; Chickasaw County, IA) | -0.053 | Mississippi | 0.125 |
| Wave 1 to Wave 2 - Worst Learners | slope | Wave 2 to Wave 3 - Worst Learners | slope |
| Group 68 (e.g. Bedford County, PA; Kenai Peninsula Borough, AK; Polk County, WI) | 0.251 | Kansas | 0.33 |
| Group 74 (e.g. Calaveras County, CA; Windham County, VT; Lumpkin County, GA) | 0.246 | Washington | 0.269 |
| Group 64 (e.g. Liberty County, FL; Johnson County, NE; Haskell County, TX) | 0.225 | Virginia | 0.259 |
| Group 18 (New Haven County, CT; Charleston County, SC; Honolulu County, HI) | 0.194 | Colorado | 0.253 |
| Group 8 (Dutchess County, NY; Fayette County, GA; El Dorado County, CA) | 0.178 | New Mexico | 0.251 |

## DISCUSSION

We estimated the county level response to the COVID-19 pandemic between subsequent waves and determined the relative importance of county demographic vs. state-level factors to predicting that response. Although we expected both state policies and county demographics to increasingly influence the response of counties to repeated waves of COVID-19 the relative importance of these two sources of variation changed over time. We found that peer county group association was most important to predicting response early in the pandemic (from wave 1-2) driven by strong patterns of learning in urban counties (Table 1, Table 2). However, state identity was more important later in the pandemic (from wave 2-3). Broadly, we found that counties which were more similar demographically or spatially were more likely to respond similarly to subsequent pandemic waves, but that the relevant axes of similarity changed over the course of the pandemic.

We found strong patterns of similarity in wave-to-wave responses at local to regional scales across the United States. Visual inspection of these patterns (Figure 2) reveals local areas and broad regions which improved their response after the first wave and held the line (the northeast US; blue/white), improved temporarily but then regressed (the mountain west, blue to green), responded badly between waves 1 and 2 but then improved (the southeast US, green to blue), and performed increasingly poorly (central Texas and the plains states; green). These patterns broadly conform to existing understanding of the course of the extent of NPI implementation and disease incidence in 2020 [19]. We also introduced new evidence on the scale at which counties tend to behave similarly and the rate at which positive correlations in response decay with distance. In particular we found that positive correlations between counties were strongest at short distances and tended to become negative at distances between 400 and 1000 km (Figure 3). Local clusters of counties are unsurprisingly similar in their response to subsequent COVID-19 waves because of similarities in state-level policies and county demographics but also because movement between neighboring counties necessarily couples the dynamics of their COVID epidemics [20, 21]. While coupling of epidemics due to proximity is still possible at intermediate distances we suggest that regional similarities in both state-level COVID policies and demographic predisposition to comply with NPIs should explain much of this remaining correlation [22]. This explanation is supported by the fact that spatial correlation patterns vary regionally with regions with smaller states, and therefore smaller county distances within states, such as the northeast crossing the zero correlation threshold more quickly with distance while regions with enormous states, such as the western US remain positively correlated over longer distances. Due to the many collinear factors interacting to drive these correlation patterns, additional analysis was required to identify probable drivers.

We, therefore, separated and identified likely mechanistic sources of county differences in response to subsequent COVID-19 waves. We predicted that the variance explained by both state and peer county group would increase during the pandemic as any uniformity in response to the early pandemic (if there was any to begin with) broke down along political and demographic lines [23, 19, 8]. Instead we found that peer county group was most important for the response between waves 1 and 2 but this importance decreased, replaced by state identity, between waves 2 and wave 3 (Figure 4, Table 1). As a result we conclude that any patterns of uniformity in response had largely broken down long before the advent of the second wave and that behavioral patterns driven by county demographics explained a substantial portion of the variation in COVID-19 case counts across the United States in that period. This pattern appears largely driven by a few county groups with strong patterns of good responses from wave 1 to wave 2 (Table 2). These groups are largely characterized by high population density urban areas which were hit hard in the first wave but mobilized to respond aggressively to the second wave. The transition from the importance of county demographics to the importance of state identity is mirrored by differences in the regional correlogram of wave responses. The difference between waves 1 and 2 is more positively correlated over very short distances and returns to positive correlations again at long distances, likely beyond state borders (Figure 3). Although states vary substantially in size across the US, the correlogram patterns match these size differences with the point at which correlations become positive again being larger in regions with larger states. Given that these positive correlations at long distances in the wave 1 to wave 2 response are unlikely to be due to state level policies, regional level factors, or spatial disease spread they likely are the result of distant counties being more similar to each other in non-spatial ways which were only relevant early in the pandemic. The increase in the importance of state identity and regional patterns between waves 2 and 3 may represent the long-term effect of divergences in COVID-19 policies between states, but it is puzzling that county demographics do not remain important, as this fracturing appears to have continued at both organizational scales [6, 24]. An alternative explanation for this pattern could be that NPI compliance behavior is driven by state and regional level reporting of COVID case counts and infection risk. This finding does not contradict the repeated finding of the enormous effect that race, socioeconomic status, and other demographic identities has on COVID-19 infection rates and outcomes [25]. Rather the county level may not be most appropriate to identify county demographic drivers of patterns of pandemic response and instead high quality finer scale data is required to understand inequities in COVID outcomes [26]. We conclude that the response between the 1st and 2nd waves at the county level was largely determined by local feasibility, acceptance, and enforcement of NPIs while the response between waves 2 and 3 was characterized by behavior based on state and regional level guidance, policies, and reporting.

We note that our analyses are largely phenomenological and provide only suggestions of possible mechanisms based on identified patterns. This type of analysis can easily break down if unknown or uncontrolled latent variables are in fact driving the associations that we have found. While a number of epidemiological factors could influence the patterns that we identified, the most likely culprit were spatio-temporal variation in seroprevalence and testing rates across the country [27, 12]. Counties with many cases may be naturally predisposed to have fewer cases in a subsequent wave simply because many susceptible individuals have been immunized by infection. For this reason we restricted our analysis to regions of the country for which we had reasonable estimates of seroprevalence with which to control our analysis [12]. While we solve the problem of confounding seroprevalence we do potentially create a new one by systematically removing regions of the country from our mixed effect model analysis. Although we urge caution in generalizing our mixed effect model findings to areas of the country outside the study area, congruence between the patterns found in the mixed effect models and those in the correlograms which span the entire continental US are encouraging. Additionally, we note that there are known to be systematic spatial biases in testing rates and case discovery rates which complicate the use of case counts for analysis [28]. While death counts are frequently used as a more reliable metric of epidemic spread [4, 29], death counts are so low in many counties during the first two waves that peak detection proved intractable. Local testing availability and demand is strongly related to a number of underlying local geographic, demographic, and socioeconomic factors which are temporally consistent, and therefore despite spatial and temporal variation in testing rates our method of comparing counties directly to themselves likely controls for much of this variation [30, 31, 32]. So despite the clear limitations of this analysis we find that it contains useful insight into the patterns and processes of response to repeated waves of COVID-19.

Additionally, we note that it is highly likely that multiple variants with different transmission rates were circulating during this period and could have contributed to some of the patterns observed. Unfortunately it is beyond the scope of this work to control for variant patterns, given the available data for this time period, but we believe our analysis largely avoids these issues for two reasons. First, there is evidence that the first identified variant of concern (alternatively B.1.1.7 or Alpha) was uncommon in the US until after our study period [33]. Second, much of the problem with spatial patterning of variants comes when directly comparing case counts in two locations at the same time. As we compare locations instead at their respective epidemic peaks within a broad date window we likely sidestep much of this concern.

Although much has been written about the relative effectiveness of various NPIs as well as the extent of implementation across the US, many gaps remain in our understanding of how humans responded to the repeated waves of COVID-19 over the past 2 years. We found that the predictors of which communities learn (or don’t learn) to deal with a pandemic tended to shift from county-level demographics to state-level policies over the course of the 2020 COVID-19 pandemic in the US. Although we focused on the pre-vaccine era of COVID-19, our findings are likely highly relevant both immediately post vaccine and especially with the rise of increasingly vaccine evasive variants, such as the omicron variant [34]. We might expect new COVID policies such as vaccination and regular antigen testing to go through a similar pattern of adoption, with early adoption being driven by county demographics and long-term trends by state-level guidance and policies. The re-implementation of familiar measures such as regular masking and social distancing for the vaccinated is an interesting hybrid policy which may show an intermediate pattern. Regardless it seems clear that the landscape of pandemic response behavior is a rapidly changing one, requiring regular reassessment of the most effective policy pressure points and the most vulnerable populations.

## Data Availability

https://doi.org/10.6084/m9.figshare.19285379

## ACKNOWLEDGMENTS

This work has been supported by the U.S. National Science Foundation RAPID grant (NSF-DEB-2031196).

## AUTHOR CONTRIBUTIONS

RLR, GF, TAD, and BDE conceived of the study. RLR performed performed analyses and wrote the first draft of the manuscript. All authors contributed subtantively to manuscript revisions.

